# Comorbidities of dementia in electronic health records: validation of a natural language processing app and comparison with data linkage

**DOI:** 10.64898/2026.10.16.26365714

**Authors:** Laiba Mirza, Rebecca Bendayan, Rob Stewart, Katrina AS Davis

## Abstract

Awareness of comorbidities in people with dementia is vital context for dementia research and clinical management. Natural Language Processing (NLP) applied to free text in the electronic health record (EHR) may help ascertain comorbidity presence. Our aim was to assess the validity of the MedCAT Comorbidity NLP app in a cohort of people with dementia.

We used text from the EHR of South London and Maudsley NHS Mental Health Trust via Clinical Records Interactive Search (CRIS), linked to structured data from local primary care (Lambeth DataNet) and hospital admission data in a cohort of 3,543 people with dementia. We evaluated MedCAT Comorbidity, examining internal validity via manual review of text and external validity by cross-validation against linked primary care and hospital data; and comparison of the predictive value of comorbidity measures for mortality.

Internal precision was highest for atrial fibrillation (100%) and lowest for diabetes (78%). Cross-validation with linked data varied substantially across conditions, with precision from 96% to 59% and recall 77% to 4%. The median number of comorbidities identified was two using any single source, or three when combined. MedCAT Comorbidity demonstrated predictive ability for mortality (HR 1.11 per condition), although the model fit was better using comorbidity from hospital admission data.

In people with dementia, the MedCAT Comorbidity app showed high precision for most conditions during internal validation. The variability of precision and recall on external validity assessment partly reflects incomplete documentation. Results from the MedCAT comorbidity app can assist researchers in understanding comorbidity of cohorts, particularly when primary care or hospital data is not available. Our findings highlight the need to develop quality assurance protocols that include internal and external validity checks before deploying general data extraction apps for specific clinical subpopulations.

**Author Summary:** People with dementia commonly have other long-term health conditions, and structured electronic health record data may not capture the whole picture. Natural language processing (NLP) may offer additional information but there is a shortage of validation studies. This study explored performance of the MedCAT NLP comorbidity app on full-text records regarding 3,543 people with dementia that had external linkage to structured data.

There were three steps of validation: firstly, internal precision against the original clinical text; secondly, compared against primary care record and hospital admission diagnosis codes; thirdly, examining the ability of comorbidity ascertainment to predict mortality. The results indicated some potential strengths and limitations of the NLP approach, which may vary by comorbidity condition. With improvements, the MedCAT comorbidity app could expand its utility in dementia research and extend into clinical scenarios. Other NLP apps may benefit from a similar validation process in clinical subgroups before deployment.

## Introduction

Dementia is well known to be associated with increased risk of death and disability, and individuals with dementia often have other physical and mental health conditions [1]. These comorbidities are important contributors to the risk of death and disability in this population, as well as accelerating decline in overall functioning and quality of life [2–4]. Therefore, research and clinical decision-making in dementia needs to account for the complex reality of living with multimorbidity, and tailor healthcare provision to these patients.

Accurate and complete data about comorbidities in people with dementia is vital. Direct surveying of people with dementia and their supporters to identify comorbidities is possible, but costly and thus limited in size and sustainability. However, there is an accumulating wealth of data derived from healthcare sources that offer efficient ascertainment at scale, with greater generalisability and inclusivity over primary data collection; but healthcare and other ‘routine’ data tends to be fragmented, collected for set reasons and recorded in different ways, leading to multiple strategies of ascertainment, each with strengths and weaknesses. Many studies leverage electronic health records (EHRs) [5,6], from which it may be possible to obtain structured diagnoses, and it may be possible to link from an EHR to registered causes of hospital admissions and deaths. However, given under-documentation of chronic conditions [7], structured data alone may be insufficient. An innovative supplementary approach would be to consider the free text entered by clinicians in EHRs, which for large-scale extractions requires natural language processing (NLP). NLP is a way of making potentially data-rich unstructured text available for analysis [8].

An NLP application developed via the Medical Concept Annotation Toolkit (MedCAT) tool [9], sought to ascertain common physical comorbidities in mental health and dementia care EHR text. MedCAT is an interface that facilitates NLP using named entity recognition and linking (NERL) allowing a semantic search from an internal thesaurus, designed to be agnostic to EHR platform and flexible in vocabulary. This derived comorbidity app was based around 21 concepts from the Unified Medical Language System (UMLS), and was refined using both supervised and unsupervised learning. A similar pipeline has been demonstrated to extract single comorbidities in people hospitalised with COVID [10], whilst the app itself was used for patients with a severe mental illness [11]. However, no study to date has applied this pipeline to clinical records of dementia patients which are likely to differ from other populations. Moreover, recent research has highlighted that data extraction pipelines may show inconsistencies when extracting for people from one sociodemographic group to another [12]. Therefore, there is a need to perform cross-validation before using this tool in people with dementia. Given the motivation for app development is the lack of easily extracted data on the subject, cross-validations are often challenging. A unique linkage between full-text data from mental health and dementia service, a local primary care EHR, and national hospitalisation data, however, presented an opportunity to triangulate results derived from the MedCAT comorbidity app and extensive structured data. Given this context, we aimed to cross-validate this application and assess its adequacy for dementia patients:

1. Assess the accuracy of selected MedCAT-derived entities in a cohort of people with dementia by comparing to the full-text (internal validity).
2. Assess the convergent validity of selected MedCAT-derived entities in this sample against the same entities from linked external data sources (external validity).
3. Describe the cohort using NLP-derived comorbidities and external-derived comorbidities, and assess whether the NLP-derived comorbidities perform as well as the external data in predicting survival (external validity).

## Methods

### Data sources

The Clinical Record Interactive Search (CRIS) platform enables researcher access to pseudonymized structured and free text data from South London and Maudsley (SLaM) Electronic Health Records (EHR) [13]. SLaM provides dementia and mental health care for around 1.3m residents of four London boroughs, and over 100 NLP algorithms have been developed over the past 15 years to extract research-relevant information from source text fields. In addition, a range of person-level data linkages have been set up between CRIS-SLaM and a local primary care database, Lambeth Data Net (LDN) [14], and with national Hospital Episode Statistics (HES) [13]. We used the CRIS data for the MedCAT NLP app.

Primary care data came from Lambeth DataNet (LDN), which covers around 405,000 people registered with a GP in Lambeth, a socially and ethnically diverse London borough [15] containing approximately a quarter of SLaM’s catchment population. All GP practices in Lambeth provide deidentified structured data from their EHRs representing more than two decades of service provision [15,14]. Conditions are extracted from the ‘journal’, where every entry is coded in triplicate with EMIS, Read and SNOMED-CT codes using internal mapping from the codes entered by clinicians.

Hospital data came from Hospital Episode Statistics (HES) Admitted Patient Care (APC), a national dataset comprising reports from English hospitals on admitted patients, discharge diagnoses and procedures as coded entities. The data are also prepared for secondary use. Each admission can include one primary diagnosis and up to 19 secondary ICD-10-coded diagnoses based on the clinical record. A recent audit concluded that HES APC data accessed for research accurately transcribe the codes recorded by hospitals [16].

### Cohort

The target population was people in Lambeth DataNet receiving a first documented diagnosis (FDD) of dementia and who were at least 65 years of age at this time. We ascertained dementia from CRIS-SLaM and LDN records using the ICD-10 and Read code lists assembled by the SAIL Dementia Cohort [17], and an algorithm that required an ICD-10 code, a strong Read code or at least two different weak Read codes for dementia, as previously deployed and described [14]. Supplementary Material file Code Lists contains this and all other code lists mentioned in this paper. We excluded patients not registered with a Lambeth GP at FDD dementia and those with no record in the CRIS-SLaM database in a window from two years before to four years after the FDD dementia. The database was extracted on 02/11/2022, although the last day represented in the hospital admission data was 31/03/2022 and the data for any patients who had opted out of secondary use of any of the constituent records were not available.

### Defining conditions

The conditions extracted from LDN and HES APC had been selected for a wider project, aiming to include those conditions particularly noteworthy for mortality casemix adjustment for people with dementia, informed by Charlson and others regarding comorbidity [20]. The code lists used for LDN were Read code lists from the CALIBER project [18], now part of HDR UK phenotype library [19]. The code lists for HES APC were ICD-10 code lists mostly drawn from the CALIBER project, while some were created by author KD. Supplementary Table S1 shows the definitions of conditions for each data source.

CRIS-SLaM free text data was processed using the MedCAT comorbidity app. Because this app had been designed separately from the project above, the choice of conditions did not exactly match the code lists. Of the 21 conditions in the app described previously [11], five conditions less known for mortality risk had no corresponding code list (migraine, chronic sinusitis, psoriasis and eczema, arthritis) were dropped. This left 16 comorbidities. Some adjustment was needed to align these: (i) merging categories – for example combining the asthma and COPD conditions from MedCAT to match the ‘Respiratory’ code lists derived from the Charlson definition; (ii) splitting categories – history of myocardial infarction was picked out from the ischaemic heart disease to match the myocardial infarction code lists. Therefore 16 MedCAT NLP conditions (17 including myocardial infarction) are compared with 12 conditions from code lists, with the overlapping list of conditions shown in Table 1. Supplementary Material Code Lists spreadsheet contains the concept list, Read and HES code lists used for defining conditions.

**Table 1.** All potential comorbidities from the MedCAT comorbidity NLP app matched to the nearest headings from ICD-10 and Read code lists (see supplementary table 2 for more detail on origin of code lists and comparability)

| MedCAT term used in CRIS-SLaM | Mapped to code list category |
| --- | --- |
| Atrial Fibrillation | Atrial fibrillation |
| Diabetes mellitus | Diabetes mellitus |
| Heart failure | Heart failure |
| Hypertension | Hypertension |
| Chronic kidney disease | Chronic renal disease |
| Cerebrovascular accident | Cerebrovascular disease |
| Transient Ischaemic Attack |  |
| Ischaemic heart diseases (except MI) | Coronary heart disease |
| Coronary arteriosclerosis |  |
| Myocardial Infarction (from IHD) | Myocardial Infarction |
| COPD | Respiratory |
| Asthma |  |
| Epilepsy | Neurological |
| Parkinson’s Disease |  |
| Multiple Sclerosis |  |
| Inflammatory bowel disease | Inflammatory bowel disease (IBD) |
| Chronic liver disease | Chronic liver disease |
| Migraine | No appropriate category |
| Chronic sinusitis |  |
| Psoriasis |  |
| Eczema |  |
| Arthritis (combined osteo and rheumatoid) |  |

COPD = Chronic Obstructive Pulmonary Disease, IHD = Ischaemic Heart Disease, MI = Myocardial infarction, TIA = Transient Ischaemic Attack

A condition mentioned in LDN or the hospital admission data was defined as pre-existing if it was recorded before the FDD of dementia. The cut-off for CRIS data for MedCAT NLP that was later than FDD of dementia was set to six months after FDD to capture those reported later in the assessment process.

### Other variables

The definition and timing of the other factors used to describe the cohort are given in Supplementary Table S2. Demographic factors included were age at diagnosis, sex, ethnicity and marital status from CRIS, and deprivation from the index of multiple deprivation based on their broad address. Dementia factors were subtype and severity at diagnosis and difficulty of activities of daily living. Finally, date of death was derived from the earliest of any death recorded in CRIS or LDN, and is assumed to be complete from previous testing against death certificates.

Missing data was present in ethnicity, marital status and dementia severity. In all cases, the missingness was considered unlikely to be at random, and an ‘unknown’ category was used to represent these.

### Ethics and data availability

CRIS-SLaM, including linkage to Lambeth DataNet and HES, has received ethical approval (Oxford Research Ethics Committee C, reference 18/SC/0372) as an anonymized data resource that does not require individual patient consent, with opt out available for individual patients. This project was approved by the CRIS oversight committee (18-109). The linked dataset and full-text extracts for internal precision were produced by the CRIS data linkage service in collaboration with the senior authors, and further extraction was possible.

The data are available at CRIS at South London and Maudsley BRC hub, but restrictions apply for confidentiality reasons, see https://www.maudsleybrc.nihr.ac.uk/facilities/clinical-record-interactive-search-cris/information-for-researchers/

### Analysis

Data cleaning was performed using R/RStudio [21], and analysis in R/RStudio with the ‘survival’ package [22] and in MS Excel.

#### Step 1: Internal precision

To evaluate the MedCAT NLP app results against manual review of source text, for each condition 50 cases were selected at random from those who had at least one annotation for that condition. Exceptions were those with less than 100 patients who were cases, where half of those cases were used, and myocardial infarction where 100 cases were used, as it had not been evaluated in the previous paper. Two metrics were used: proportion of annotations judged correct; proportion of patients selected judged to have the condition. These reflect the instance- and case-level precision.

#### Step 2: Cross-validation with external resources

Output from the CRIS-SLaM MedCAT NLP comorbidity app was compared against LDN and the hospital admission data, extracted by code lists based on previous lists produced by the CALIBER study. The combined LDN and the hospital admission data (i.e. positive in either source) was used as a ‘gold standard’ for the sake of defining precision and recall.

#### Step 3: Further external validity

A descriptive analysis was carried out of different definitions of pre-existing comorbidity, including the most prevalent conditions in each source, the overall number of comorbidities according to each source, and the relative contribution of each source to the overall prevalence. For the latter we considered the group of people with dementia that could be identified as having a particular comorbid condition by any of MedCAT NLP, LDN or the hospital admission data, then calculated the proportion identified by MedCAT NLP, those included by adding LDN ascertainment, and finally those added through the hospital admission data ascertainment.

Further external validation was carried out by comparing data derived from the different sources with respect to prediction of mortality in the four years after the index dementia diagnosis. The assumption was that comorbidities should be independent risk-factors for mortality, and therefore the better the fit of the model using comorbidity ascertainment, after adjustment (for age at diagnosis, sex, dementia severity at diagnosis and dementia type), the more useful that method of ascertainment in dementia research. We evaluated the performance of models based on different sources and combinations by performing a survival analysis with Cox proportional hazards regression, with comorbidity and confounders as the independent variables, and mortality as the outcome. Assumption of proportional hazards was assessed using Schoenfeld residuals.

The standard test for comparing Cox models, the log likelihood ratio test, is indicated for comparing nested Cox models. The NLP and linkage models are not nested, so we considered multiple statistics: the strength of association of conditions with mortality (hazard ratio, HR), the log likelihood ratio test (LRT) of each model against no model, and the Akaike Information Criterion (AIC) [23]. A hazard ratio (HR) ≥1.2 was used to define a risk factor, and high statistical significance was defined as p(z)<0.001. Lower AIC values and higher LRT values indicate better model fit, provided the cohort is the same in the models compared.

## Results

### Cohort

The initial cohort comprised 4,161 individuals with a documented diagnosis of dementia who were registered with a Lambeth GP. After excluding 618 individuals who never received care from SLaM, 3,543 individuals remained (Supplementary Table S3). Participants were predominantly (63%) above 80 years or above at diagnosis. Men accounted for 41% of the cohort. The population was ethnically diverse, comprising 58% White, 26% Black and 5.9% Asian participants. Nearly half of the cohort (49%) lived in the most deprived tertile nationally, and only 25% were married or cohabiting. Alzheimer’s disease was the commonest subtype (63%), followed by vascular or mixed dementia (18%). Dementia severity at diagnosis was 37% severe, compared with 21% moderate and 27% mild disease.

MedCAT NLP identified hypertension (64%), diabetes (42%) and stroke/TIA (27%) as the most common comorbidities, shown in Table 2. Overall, patients had a median of two comorbidities (IQR 1–3; mean 1.88) from the twelve conditions studied.

**Table 2.**
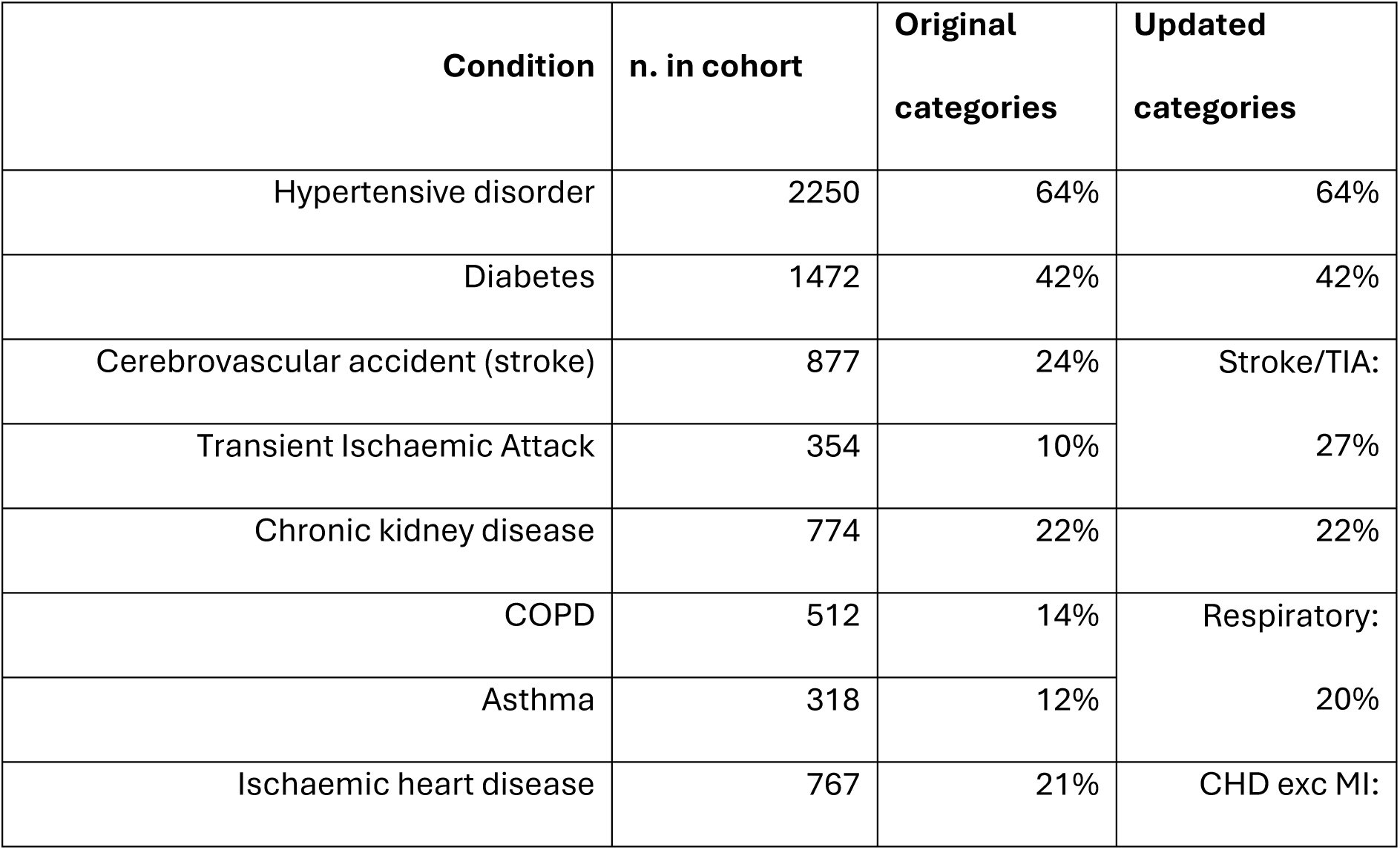
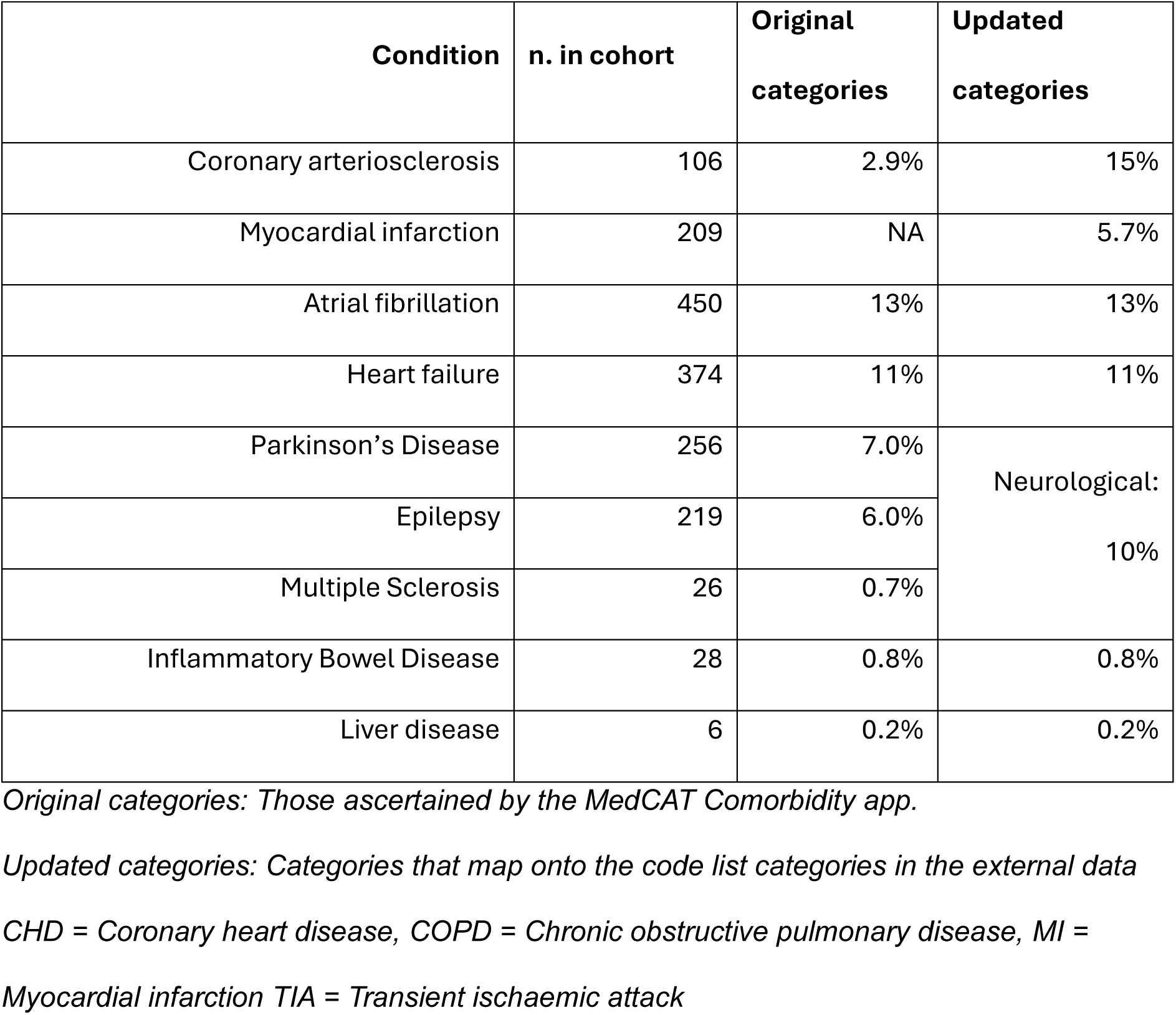
The prevalence of pre-existing comorbidities in a dementia cohort identified by MedCAT NLP on mental health and dementia care electronic health records.

| Condition | n. in cohort | Original categories | Updated categories |
| --- | --- | --- | --- |
| Hypertensive disorder | 2250 | 64% | 64% |
| Diabetes | 1472 | 42% | 42% |
| Cerebrovascular accident (stroke) | 877 | 24% | Stroke/TIA:<br>27% |
| Transient Ischaemic Attack | 354 | 10% |  |
| Chronic kidney disease | 774 | 22% | 22% |
| COPD | 512 | 14% | Respiratory:<br>20% |
| Asthma | 318 | 12% |  |
| Ischaemic heart disease | 767 | 21% | CHD exc MI: |
| Coronary arteriosclerosis | 106 | 2.9% | 15% |
| Myocardial infarction | 209 | NA | 5.7% |
| Atrial fibrillation | 450 | 13% | 13% |
| Heart failure | 374 | 11% | 11% |
| Parkinson's Disease | 256 | 7.0% | Neurological:<br>10% |
| Epilepsy | 219 | 6.0% |  |
| Multiple Sclerosis | 26 | 0.7% |  |
| Inflammatory Bowel Disease | 28 | 0.8% | 0.8% |
| Liver disease | 6 | 0.2% | 0.2% |

*Original categories: Those ascertained by the MedCAT Comorbidity app.*
*Updated categories: Categories that map onto the code list categories in the external data*
*CHD = Coronary heart disease, COPD = Chronic obstructive pulmonary disease, MI = Myocardial infarction TIA = Transient ischaemic attack*

#### Step 1: Internal precision

Most conditions had more than 200 instances of annotation and over 100 patients flagged, except for chronic liver disease, multiple sclerosis and IBD. Table 3 shows that precision at instance level was over 90% for 11 of 17 conditions, while diabetes had the lowest precision at 84%, and ischaemic heart disease and heart failure both had 89%. Precision at a patient level was over 90% for four of 17 conditions, and a further eight were above 80%, while TIA had the lowest patient-level precision at 76%, diabetes was 78%, and epilepsy was 77%. Reasons for false positive annotations in dementia patients are shown in Supplementary Table S4.

**Table 3.** Detail of internal validation of MedCAT comorbidities against manual annotation in the dementia cohort (n=3,543)

| Condition | Instance level |  |  | Patient level (1+ instance) |  |  |
| --- | --- | --- | --- | --- | --- | --- |
|  | n. in CRIS | examined | precision | n. in CRIS | examined | precision |
| Diabetes | 25786 | 349 | 84% | 1770 | 50 | 78% |
| Hypertensive disorder | 13830 | 199 | 95% | 2418 | 50 | 96% |
| Chronic kidney disease | 3470 | 231 | 98% | 846 | 50 | 98% |
| Heart failure | 2500 | 150 | 89% | 472 | 50 | 84% |
| Atrial fibrillation | 1815 | 213 | 99% | 459 | 50 | 100% |
| Inflammatory Bowel Disease | 128 | 61 | 88% | 40 | 20 | 80% |
| Liver disease | 23 | 14 | 100% | 11 | 6 | 100% |
| Cerebrovascular accident | 12079 | 343 | 91% | 1426 | 50 | 86% |
| Transient Ischaemic Attack | 1731 | 170 | 90% | 465 | 50 | 76% |
| Parkinson's Disease | 4517 | 259 | 86% | 340 | 50 | 81% |
| Epilepsy | 1756 | 189 | 86% | 345 | 50 | 77% |
| Multiple Sclerosis | 65 | 32 | 94% | 24 | 12 | 80% |
| COPD | 3515 | 335 | 94% | 510 | 50 | 84% |
| Asthma | 2329 | 243 | 92% | 479 | 50 | 92% |
| Ischaemic heart disease | 3565 | 215 | 89% | 816 | 50 | 88% |
| Myocardial infarction | 696 | 333 | 99% | 209 | 100 | 99% |
| Coronary arteriosclerosis | 261 | 156 | 92% | 104 | 50 | 80% |

Condition = concept the NLP flagged.
Examined = all instances in 50 random patients from those flagged by the NLP in the cohort, or if less than 50 cases, a random 50% of cases. MI was greater, as this had not been previously evaluated for any cohort.
Instance level = flags from NLP found in the free text of CRIS, where precision is proportion matching manual annotation (true mention of current condition in the patient)
Patient level = patients with at least one instance, where precision is proportion where condition was present in the patient according to manual annotation of all flagged instances in the patient (at least one true mention of current condition in the patient)

#### Step 2: Cross-validity (external validity)

Table 4 shows the comparison between the MedCAT NLP and codelist extraction from external sources (primary care and hospital admission data) of pre-existing condition, ordered by prevalence by MedCAT NLP. In all conditions except diabetes, more cases were found by the external sources than by the MedCAT NLP. Precision estimates against the external source ranged from 96% to 59% with hypertension, renal and atrial fibrillation having precision over 90% but coronary heart disease, neurological and chronic liver disease having precision less than 70%. Recall estimates for MedCAT NLP were heterogeneous, with hypertension and diabetes having a recall of over 70%, renal, respiratory and coronary heart disease 70 to 50%, and six further conditions between 50 and 30%, including myocardial infarction at 32%. Chronic liver disease was an outlier at 4%.

**Table 4.** Conditions ascertained from mental healthcare text via the MedCAT NLP comorbidity app mapped on to conditions ascertained from linked external data (ICD-10 codes from hospitalisations and Read codes from primary care) to calculate precision and recall (n=3543 with a diagnosis of dementia)

| Condition (updated categories) | n. found by MedCAT in text | n. found by linkage | Precision of comorbidity app | Recall of comorbidity app |
| --- | --- | --- | --- | --- |
| Hypertension | 2250 | 2792 | 95% | 77% |
| Diabetes | 1472 | 1459 | 80% | 73% |
| Stroke / TIA | 941 | 1580 | 79% | 47% |
| Renal | 774 | 1320 | 90% | 51% |
| Respiratory | 725 | 1047 | 86% | 56% |

| **Condition (updated categories)** | n. found by<br>MedCAT in<br>text | n. found by<br>linkage | Precision of<br>comorbidity<br>app | Recall of<br>comorbidity<br>app |
| --- | --- | --- | --- | --- |
| Coronary heart disease | 514 | 581 | 61% | 54% |
| Atrial Fibrillation | 450 | 1028 | 96% | 42% |
| Heart Failure | 374 | 788 | 71% | 41% |
| Myocardial Infarction | 209 | 505 | 79% | 32% |
| Neurological | 247 | 282 | 59% | 34% |
| Inflammatory bowel disease | 21 | 47 | 84% | 47% |
| Chronic liver disease | 5 | 67 | 65% | 4% |

TIA = Transient Ischaemic Attack

#### Analysis 3: Further external validity

Comparing the description of cohort comorbidity: Supplementary Table S5 shows the most common conditions are the same, regardless of data source, with four conditions being in the top 5 for all (hypertension, diabetes, stroke/TIA and respiratory), with renal being the fifth in MedCAT NLP and primary care data, while AF was more common in hospital inpatient data. All individual sources extracted a median of 2 (IQR 1-3) conditions per person. Combining all three sources increased this, resulting in a median of 3 (IQR 2-4) conditions.

Of all cases of comorbidity that could have been identified in this cohort from any information source, the MedCAT NLP app identified around 53%. Fig 1 breaks this down by condition, and also shows the contribution of hospital admission data (particularly for stroke/TIA, atrial fibrillation, heart failure and MI) and primary care data (particularly for renal, IBD and chronic liver disease).

**Fig 1.**
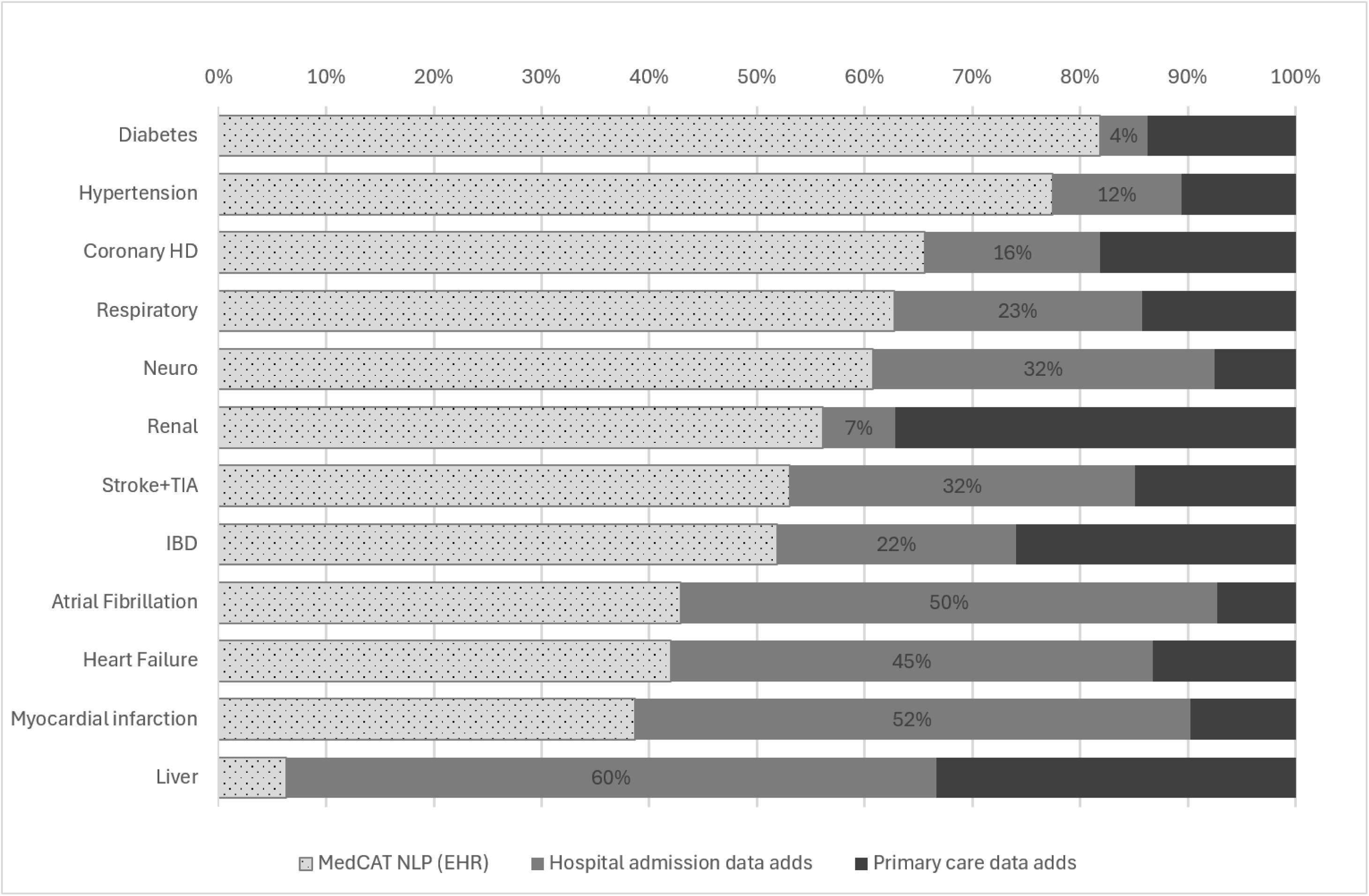
Contribution of the three data sources to the total patients identified with one of the listed comorbidities, with proportion of cases hospital admissions (HES) adds to MedCAT ascertainment labelled. Coronary HD = Coronary Heart Disease, Neuro = Neurological, TIA = Transient Ischaemic Attack

### Modelling comorbidities and mortality

During the four-year follow-up, mortality in the dementia patient cohort was 43%. Table 5 shows effect of comorbidities on the risk of mortality, comparing Cox survival models using the three data sources on whether there were significant conditions, and model fit through AIC and LRT. The MedCAT NLP-derived comorbidity model found five conditions with hazard ratios (HR) ³1.2, but none met the significance target. For primary care data, two significant risk factors were found, and for hospital admission data, three significant risk factors. Combining all three sources gave four significant risk factors. Model statistics were most favourable (AIC minimised, LRT maximised) for hospital codes, which was slightly better than combined data. Cox models using number of comorbidities rather than individual comorbidities, found this to be a highly significant risk factor for mortality in all sources. HRs per condition ranged from 1.11 with the MedCAT NLP data, to 1.21 with hospital data. LRT was maximised with hospital data, although AIC was slightly lower for the primary care codes.

**Table 5.** Results of Cox regression for mortality including either individual comorbidities (upper) or number of comorbidities (lower) when considering ascertainment from three sources – MedCAT-NLP (from specialist mental health & dementia care data, CRIS), Primary care data (from Lambeth DataNet) via codelist, Hospital admission data (from Hospital Episode Statistics (HES-APC)) via codelist, or combined (any of the above).

|  | Data Source |  |  |  |  |  |  |  |
| --- | --- | --- | --- | --- | --- | --- | --- | --- |
|  | MedCAT-NLP |  | Primary care data |  | Hospital admission data |  | Any of MedCAT-NLP, primary or admission |  |
|  | HR | p-value | HR | p-value | HR | p-value | HR | p-value |
| **Hypertension** | 0.90 | 0.065 | 0.85 | 0.002 | 1.08 | 0.223 | 0.96 | 0.497 |
| **Diabetes** | 0.97 | 0.569 | 1.02 | 0.684 | 1.08 | 0.191 | 0.94 | 0.244 |
| **Stroke/TIA** | 1.09 | 0.213 | 1.09 | 0.205 | 1.14 | 0.018 | 1.23 | <0.001 |
| **Renal** | 1.18 | 0.020 | 1.22 | 0.001 | 1.26 | 0.002 | 1.20 | 0.002 |
| **Respiratory** | 1.21 | 0.005 | 1.29 | <0.001 | 1.34 | <0.001 | 1.31 | <0.001 |
| **AF** | 1.31 | 0.001 | 1.22 | 0.005 | 1.33 | <0.001 | 1.32 | <0.001 |
| **Heart Failure** | 1.37 | 0.001 | 1.61 | <0.001 | 1.50 | <0.001 | 1.34 | <0.001 |
| **Coronary HD** | 1.15 | 0.086 | 0.93 | 0.419 | 0.91 | 0.260 | 0.97 | 0.707 |
| **Neuro** | 1.22 | 0.050 | 1.35 | 0.010 | 1.29 | 0.009 | 1.23 | 0.012 |
| **MI** | 1.09 | 0.480 | 1.16 | 0.111 | 1.06 | 0.538 | 1.16 | 0.065 |
| **Liver** | 1.97 | 0.243 | 1.19 | 0.439 | 1.11 | 0.645 | 1.09 | 0.633 |
| **IBD** | 0.97 | 0.930 | 1.07 | 0.800 | 0.95 | 0.850 | 0.94 | 0.775 |
| *LRT (model stats):* | 597 |  | 635 |  | **704** |  | 675 |  |
| *AIC (model stats):* | 23486 |  | 23447 |  | **23379** |  | 23407 |  |
|  | MedCAT-NLP |  | Primary care data |  | Hospital admission data |  | Any of MedCAT-NLP, primary or admission |  |
| **N. (HR per comorbidity)** | 1.11 | <0.001 | 1.16 | <0.001 | 1.21 | <0.001 | 1.19 | <0.001 |
| *LRT (model stats):* | 561 |  | 569 |  | **664** |  | 613 |  |
| *AIC (model stats):* | 23498 |  | **23391** |  | 23491 |  | 23434 |  |

Footnotes: Cox regression for each source included all comorbidities (upper) or number of comorbidities (lower) and age at diagnosis, sex, dementia severity at diagnosis and dementia type. Shading highlights items meeting pre-specified target levels, while model statistics in bold are the optimal.
AF = Atrial Fibrillation, Coronary HD = Coronary Heart Disease, MI = Myocardial Infarction, TIA = Transient Ischaemic Attack
HR = Hazard Ratio, LRT = Likelihood Ratio Test, AIC = Akaike Information Criterion

## Discussion

This study aimed to assess the validity of an app that combined a suite of NLP algorithms to extract mentions of comorbid conditions from EHR free text for the task of ascertaining pre-morbid comorbidities in people with dementia. MedCAT is shown to be versatile across different clinical settings, but there is a need to assess performance when applying any phenotyping algorithm to new contexts [9,24]. The context of dementia is important, because symptoms, functioning and needs in dementia are often related to both dementia and comorbidities, and more research is important [1,25]. We designed and implemented a three-step assessment strategy which involved: i) internal validity against source text, ii) cross-validation against alternative data sources, and iii) comparative performance in modelling (external validity). This was achieved through a unique dataset that linked mental health EHR (CRIS) to linked data from primary health care services (LDN), and to hospital admission data (HES). Overall, the results showed that the app has high levels of precision for most conditions, but there is room for improvement in some conditions, particularly diabetes, and other data could have better performance for case-finding and mortality modelling, if available.

Internal validity checks showed that most instances flagged were accurate, with only three conditions having a precision below 90%, and two of these had specific boundary issues (i.e. how to label pre-diabetes and resolved symptoms of coronary heart disease). The mention of disorders in non-specific text in the notes gave rise to error – commonly, for example, from blood results for HbA1c accompanied by text referring to the reference range for diabetes which were incorrectly being flagged as the presence of the condition. Although this source text was stereotyped, it differed from different labs and over time, making it difficult to filter out with simple pattern-matching.

Cross-validation of NLP apps is important, because good performance of the app against the source material cannot assess the adequacy of the source material, and it is the combination that is important for the utility of the NLP output [8,26]. The external validity of the comorbidity app was examined against two linked data sources, primary care and hospital, extracted using code lists selected for case-mix adjustment for mortality in dementia. Like designing NLP pipelines, building code lists is not straightforward or standardised; both need clear definition of what is to be extracted and a realistic attitude to trade-offs in time and different aspects of accuracy. This had motivated our use of pre-defined code lists. However, in using the MedCAT definitions of comorbidity alongside the CALIBER definitions of comorbidity, aligning definitions may have been imprecise and poor alignment may lead to relatively poorer precision on cross-validation [27], such as for coronary heart disease (88% internal, 61% external).

Recall of the NLP tool against the linked data was quite variable, from 77% for hypertension to 32% for MI, with a mean of 56%. This incomplete ascertainment of comorbidities remains an important limitation. The observed patterns of missing information are unlikely to be random and reflect both the accuracy achieved by the NLP algorithms to ascertain recorded mentions, and how likely the condition was to be recorded in the mental healthcare EHR in the first place. Recording will be influenced by disease factors (chronicity, severity, intrusiveness), and healthcare factors (documentation practices, clinician interest, considerations for medication) [7]. The effect of under-ascertainment, both overall and between different conditions, will need to be assessed when such data are used for future research or clinical purposes, for instance to prevent bias in recording becoming embedded in research [28].

Linked data can contribute substantially to the ascertainment of comorbidity, including hospital data on acute cases of atrial fibrillation, heart failure and myocardial infarction, and primary care data for ascertaining chronic respiratory, renal, diabetes and hypertension conditions. Overall, adding in hospital and primary care data to the NLP data increases the number of conditions ascertained from two to three. However, it is not clear whether this extra sensitivity is an advantage for onward research, as it may lead to the inclusion of conditions that have little effect on future morbidity and mortality. We further investigated the external validity of the app by examining its value in predicting mortality, comparing the MedCAT app with primary care codes, hospital admission codes and the combined data. The comorbidities studied were mainly in the Charlson comorbidity tool, which was designed to control for the effect of comorbidities on mortality, and the model used was based on the original study [20], so we would expect these conditions to collectively predict mortality, and many individually too. The NLP data appeared to be useful for adjustment of health, but was improved by adding hospital data and primary care data. Hospital data gave the best fit. This is consistent with previous evidence that hospital-derived comorbidity data often outperforms community-derived data for mortality prediction [29]. This may be because hospital admission data is only present if the patient was sufficiently sick to need hospital, which has prognostic importance in itself.

Using NLP apps to ascertain patient history, including comorbidities, in real-time through the EHR interface could save time in routine clinical practice, and could offer extra value by feeding into recommendations for evidence-based actions [30]. The performance of the MedCAT comorbidity app for people with dementia is unlikely to be sufficient for patient-level clinical use at this point, mainly due to lower precision on some highly important conditions such as diabetes. Our findings support the view that NLP systems require internal and external validation before being deployed in clinical services, and that this needs to involve both the extraction algorithm but also documentation practices [31]. Further development would bring the app up to an acceptable standard of reflecting the data in the EHR, but that EHR remaining in a data silo may leave vital gaps in the clinical picture in a condition as multifaceted as dementia [32]. Live combination of data from the multiple healthcare providers that patients interact with would address the information gaps demonstrated here. The main impediments to using data-sharing in patient care seem to be governance and acceptability, rather than technological [33].

### Strengths and limitations of the study

A major strength of this study is its use of a unique large linked dataset that brings together primary care, hospital episodes, and specialist mental health care records. This is a powerful resource for dementia research in the UK to capture the complex clinical needs of these patients, which includes brain and body. Part of the depth of information is the inclusion of free-text records, and the application of natural language processing (NLP), including through MedCAT, enabling the extraction of clinically relevant information that is often overlooked or not available in other datasets [34]. For this study, the breadth of data made it possible to evaluate the MedCAT comorbidity app at both the source data level and by cross-reference with external sources. This validation can guide adoption of this app for dementia patients and allow further research to improve and utilise NLP tools in wider cohorts. We suggest that a systematic examination of internal and external validity, similar to that outlined here, should be standard before NLP apps are used in specific clinical populations.

There are, however, some important limitations. Firstly, the NLP specification and codelist specification were separate, rather than using an integrative process, potentially affecting the cross-validation results for some conditions. The nature of routinely collected data, including the retrospective nature and under-documentation, may have introduced misclassification and subsequent bias. Lastly, the study was conducted in a single geographical area, albeit one of the most ethnically diverse areas in the UK, which may limit generalisability to other populations, and the relatively low prevalence of some conditions in our cohort leads to uncertainty about the tool’s precision and recall for those conditions.

## Conclusion

This study illustrates that NLP-based extraction of comorbidities from free-text mental health & dementia care EHRs can make a meaningful contribution to dementia research, enriching the record by making the vast amount of data in full-text accessible. Moreover, this study highlights the need to establish validation protocols for these types of apps in the populations where it will be applied and exemplifies a systematic way to assess internal and external validity. The incomplete ascertainment of comorbidities remains an important consideration, with a contributing factor being underlying gaps in the medical history from a single source. This suggests that strategies such as supplementing NLP outputs with data from structured sources may be beneficial for some uses, for example, primary care records will capture more chronic ongoing problems whereas hospital records capture acute conditions requiring inpatient care. This NLP app, alongside refinements informed by this study, has great potential for supporting patient care for people with dementia. Further evaluation should ensure that this application and other technology implemented in electronic health records is both accurate and valuable, saving clinicians time and better able to care for patients. Therefore, our findings have implications for NLP developers, data scientists, clinicians, healthcare organisations and patients.

## Supporting information

Code List Spreadsheet

## Funding

This study/research is funded by the National Institute for Health and Care Research (NIHR) Biomedical Research Centre (BRC): Maudsley. The views expressed are those of the author(s) and not necessarily those of the NIHR or the Department of Health and Social Care.

## Declaration of Conflicts of Interest

None

## Research data transparency and availability

The data used in this study are available at CRIS at South London and Maudsley BRC hub, but restrictions apply for data confidentiality, see

https://www.maudsleybrc.nihr.ac.uk/facilities/clinical-record-interactive-search-cris/information-for-researchers/

The authors have completed a RECORD / STROBE checklist, available as Supplementary Table S6.

## Supporting information captions

**S1. Table.** Definitions of conditions for each data source.

**S2. Table.** Definitions and timing of other factors used to describe the cohort.

**S3. Table.** Cohort derivation and characteristics.

**S4. Table.** Reasons for false-positive MedCAT NLP annotations in people with dementia.

**S5. Table.** Comparison of the most common comorbidities across data sources.

**Supplementary Material Code List Spreadsheet.** Code lists used to define dementia and comorbid conditions.

## Notes

### Competing Interest Statement

The authors have declared no competing interest.

### Author Declarations

Oxford Research Ethics Committee C of NHS Health Research Authority gave ethical approval to Clinical Records Information Search at South London and Maudsley (reference 18/SC/0372). Oversight committee at Clinical Records Information search gave ethical approval for this work.

## References

1. Stirland LE, Choate R, Zanwar PP, Zhang P, Watermeyer TJ, Valletta M, et al. Multimorbidity in dementia: current perspectives and future challenges. Alzheimers Dement. 2025;21(8):e70546.

2. Garre-Olmo J, Ponjoan A, Inoriza JM, Blanch J, Sanchez-Perez I, Cubi R, et al. Survival, effect measures, and impact numbers after dementia diagnosis: a matched cohort study. Clin Epidemiol. 2019;11:525–542.

3. Haaksma ML, Rizzuto D, Ramakers I, Garcia-Ptacek S, Marengoni A, van der Flier WM, et al. The impact of frailty and comorbidity on institutionalization and mortality in persons with dementia: a prospective cohort study. J Am Med Dir Assoc. 2019;20(2):165–170.e2.

4. Livingston G, Huntley J, Liu KY, Costafreda SG, Selbaek G, Alladi S, et al. Dementia prevention, intervention, and care: 2024 report of the Lancet standing Commission. Lancet. 2024;404(10452):572–628.

5. Perera G, Soremekun M, Breen G, Stewart R. The psychiatric case register: noble past, challenging present, but exciting future. Br J Psychiatry. 2009;195(3):191–193.

6. Davis KA, Farooq S, Hayes JF, John A, Lee W, MacCabe JH, et al. Pharmacoepidemiology research: delivering evidence about drug safety and effectiveness in mental health. Lancet Psychiatry. 2019.

7. Hersh WR, Weiner MG, Embi PJ, Logan JR, Payne PRO, Bernstam EV, et al. Caveats for the use of operational electronic health record data in comparative effectiveness research. Med Care. 2013;51:S30–S37.

8. Velupillai S, Suominen H, Liakata M, Roberts A, Shah AD, Morley K, et al. Using clinical natural language processing for health outcomes research: overview and actionable suggestions for future advances. J Biomed Inform. 2018;88:11–19.

9. Kraljevic Z, Searle T, Shek A, Roguski L, Noor K, Bean D, et al. Multi-domain clinical natural language processing with MedCAT: the medical concept annotation toolkit. Artif Intell Med. 2021;117:102083.

10. Carr E, Bendayan R, Bean D, Stammers M, Wang W, Zhang H, et al. Evaluation and improvement of the National Early Warning Score (NEWS2) for COVID-19: a multi-hospital study. BMC Med. 2021;19(1):23.

11. Bendayan R, Kraljevic Z, Shaari S, Das-Munshi J, Leipold L, Chaturvedi J, et al. Mapping multimorbidity in individuals with schizophrenia and bipolar disorders: evidence from the South London and Maudsley NHS Foundation Trust Biomedical Research Centre case register. BMJ Open. 2022;12(1):e054414.

12. Omar M, Soffer S, Agbareia R, Bragazzi NL, Apakama DU, Horowitz CR, et al. Socio-demographic biases in medical decision-making by large language models: a large-scale multi-model analysis. medRxiv [Preprint]. 2024:2024.10.29.24316368.

13. Perera G, Broadbent M, Callard F, Chang CK, Downs J, Dutta R, et al. Cohort profile of the South London and Maudsley NHS Foundation Trust Biomedical Research Centre case register: current status and recent enhancement of an electronic mental health record-derived data resource. BMJ Open. 2016;6(3):e008721.

14. Davis KAS, Mueller C, Ashworth M, Broadbent M, Jewell A, Molokhia M, et al. What gets recorded, counts: dementia recording in primary care compared with a specialist database. Age Ageing. 2021;50(6):2206–2213.

15. Ashworth M, Durbaba S, Whitney D, Crompton J, Wright M, Dodhia H. Journey to multimorbidity: longitudinal analysis exploring cardiovascular risk factors and sociodemographic determinants in an urban setting. BMJ Open. 2019;9(12):e031649.

16. Murray ML, Pinches H, Mafham M, Hartley S, Carpenter J, Landray MJ, et al. Use of NHS Digital datasets as trial data in the UK: a position paper. 2022.

17. Schnier C, Wilkinson T, Orton C, North L, Rochford R, Sudlow C. The Secure Anonymised Information Linkage databank Dementia e-cohort (SAIL-DeC). Int J Popul Data Sci. 2019;4(3).

18. Denaxas S, Gonzalez-Izquierdo A, Direk K, Fitzpatrick NK, Fatemifar G, Banerjee A, et al. UK phenomics platform for developing and validating electronic health record phenotypes: CALIBER. J Am Med Inform Assoc. 2019;26(12):1545–1559.

19. HDR UK. Phenotype Library [Internet]. 2026. Available from: https://phenotypes.healthdatagateway.org/.

20. Charlson ME, Pompei P, Ales KL, MacKenzie CR. A new method of classifying prognostic comorbidity in longitudinal studies: development and validation. J Chronic Dis. 1987;40(5):373–383.

21. R Core Team. R: A language and environment for statistical computing. Vienna: R Foundation for Statistical Computing; 2024.

22. Therneau T. A package for survival analysis in R [Internet]. 2024. Available from: https://cran.r-project.org/.

23. GraphPad. Comparing Cox proportional hazards regression models [Internet]. 2024. Available from: https://www.graphpad.com/.

24. Brandt PS, Pacheco JA, Adekkanattu P, Sholle ET, Abedian S, Stone DJ, et al. Design and validation of a FHIR-based EHR-driven phenotyping toolbox. J Am Med Inform Assoc. 2022;29(9):1449–1460.

25. Swain S, Kantilal K, Suryadevara V, Stirland LE. Multimorbidity research in dementia: it’s time to shift to a person-centred approach. Front Dement. 2026;5.

26. Fu S, Wang L, Moon S, Zong N, He H, Pejaver V, et al. Recommended practices and ethical considerations for natural language processing-assisted observational research: a scoping review. Clin Transl Sci. 2023;16(3):398–411.

27. Denaxas S, Parkinson H, Fitzpatrick N, Sudlow C, Hemingway H. Analyzing the heterogeneity of rule-based EHR phenotyping algorithms in CALIBER and the UK Biobank. bioRxiv [Preprint]. 2019:685156.

28. Smith L, Simpson G, Singh N, Wells L, Dambha-Miller H. Using electronic health records to understand multimorbidity in older people: a scoping review. Eur Geriatr Med. 2025;16(4):1163–1206.

29. Quan H, Li B, Couris CM, Fushimi K, Graham P, Hider P, et al. Updating and validating the Charlson Comorbidity Index and score for risk adjustment in hospital discharge abstracts using data from 6 countries. Am J Epidemiol. 2011;173(6):676–682.

30. Harland R, Wang T, Codling D, Polling C, Broadbent M, Newton H, et al. Developing clinical informatics to support direct care and population health management: the VIEWER story. BMJ Health Care Inform. 2025;32(1).

31. Au Yeung J, Shek A, Searle T, Kraljevic Z, Dinu V, Ratas M, et al. Natural language processing data services for healthcare providers. BMC Med Inform Decis Mak. 2024;24(1):356.

32. Hazan J, Howard R, Isaacs JD. England’s dementia diagnosis landscape: strengths, limitations, and future directions. Int J Geriatr Psychiatry. 2026;41(7):e70236.

33. Understanding Patient Data. Long Read: The Single Patient Record: what the legislation does - and what it will take to deliver it [Internet]. 2026. Available from: https://www.understandingpatientdata.org.uk/long-read-single-patient-record-what-legislation-does-and-what-it-will-take-deliver-it.

34. NIHR Maudsley Biomedical Research Centre. Natural Language Processing (NLP) Service [Internet]. 2024. Available from: https://www.maudsleybrc.nihr.ac.uk/facilities/clinical-record-interactive-search-cris/cris-natural-language-processing/.

