## Supplementary Materials for "Comorbidities of dementia in electronic health records: validation of a natural language processing app and comparison with data linkage"

Supplementary Materials Code List (see separate excel document)

S1 Supplementary Table 1: Extra information about comparability of modes of identification

| **Heading** | **Read code list**  **used in primary care data (LDN)** | **ICD-10 list**  **used in hospital admission data (HES)** | **MedCAT term**  **used in CRIS-SLaM** | **Notes** |
| --- | --- | --- | --- | --- |
| Atrial fibrillation | CALIBER list: af | Own | Atrial Fibrillation | Nil |
| Cerebrovascular disease  (TIA / stroke) | CALIBER tia_gprd + stroke_nos_gprd + ischaemic_stroke_gprd + haem_stroke_gprd | CALIBER tia_hes + stroke_nos_hes + ischaemic_stroke_hes + haem_stroke_hes | Combined: TIA, Cerebrovascular accident | Wider than ischaemic stroke, containing mini-strokes (TIA) and haemorrhagic stroke |
| Coronary heart disease (IHD/angina) | CALIBER list: angina and uangina | CALIBER list: angina and uangina | Combined: IHD, Coronary arteriosclerosis | Code lists refer to angina, whereas MedCAT is any mention of coronary or IHD (including anginas). Original MedCAT included MI, but extracted as separate for this study |
| Diabetes | CALIBER dm_gprd | CALIBER dm_hes | Diabetes | No explicit differentiation between types, severity or treatment type |
| Heart failure | CALIBER hf_gprd | CALIBER hf_hes | Heart failure | CALIBER is chronic only, MedCAT does not distinguish. All include ‘congestive’ and other types |
| Hypertension | CALIBER list: ht, bp and bp_meds | Own | Hypertensive disorder | No differentiation between treated, untreated or refractory |
| Inflammatory bowel disease (IBD) | CALIBER list: ibd_gprd | CALIBER list: ibd_hes | IBD | Nil |
| Liver disease, chronic | CALIBER liver_charlson_gprd (3,4) | CALIBER liver_charlson_hes (3,4) | Chronic liver disease | No explicit differentiation between types or severity |
| Myocardial infarction (MI) | CALIBER myo_infarct_gprd | CALIBER myo_infarct_hes | MI | Original MedCAT included other IHD, but extracted as separate for this study |
| Neurological | Derived from epilepsy, Parkinsons and MS CALIBER lists | Derived from epilepsy, Parkinsons and MS CALIBER lists | Combined: epilepsy, Parkinsons, MS | Nil |
| Renal disease, chronic | CALIBER renal_gprd (5,6,7) | CALIBER renal_hes (5,6,7) | Chronic kidney disease | No explicit differentiation between severity |
| Respiratory disorders, chronic | CALIBER respiratory_charlson_gprd | CALIBER respiratory_charlson_hes | Combined: COPD, asthma | MedCAT is more likely to identify episodic rather than permanent morbidity |

Notes: HES = Hospital Episode Statistics (Admitted), LDN = Lambeth DataNet, MedCAT = Medical Concept Annotation Tool. CALIBER is now available via the HDR-UK phenotyping portal. “Charlson” refers to CALIBER list designed to extract disorder for the Charlson Comorbidity Index.

COPD = Chronic Obstructive Pulmonary Disease, IBD = Inflammatory Bowel Disease, IHD = Ischaemic Heart Disease, TIA = Transient Ischaemic Attach

S2 Supplementary Table 2 Variables used in analysis, with index being the first documented dementia diagnosis date

| **Variable** | **Source** | **Timing** | **Detail** |
| --- | --- | --- | --- |
| Basic |  |  |  |
| Sex | LDN – Primary care data | NA | Female / Male |
| Age (at diagnosis) | derived | index | From year of birth from LDN (approximate as year of birth, not date of birth given) Derived from year of birth recorded in primary care data” |
| Dementia |  |  |  |
| Subtype of dementia | CRIS/LDN -Mental healthcare full text/ primary care data | see detail | Taking CRIS most recent diagnosis if present Taking the most recent diagnosis recorded in mental healthcare full text, and if most frequently recorded dementia subtype in primary care data” no CRIS diagnosis: AD / Vascular or mixed AD and vascular dementia / Other / Unspecified |
| Dementia severity | CRIS Mental healthcare full text | closest to index (within 6 months) | Severity from MMSE if there was a valid MMSE in the six month window (**valid** = has denominator >26 **and** is not 0). If no valid MMSE in window, use HONOS cognitive score if this is in six month window.  MMSE was used in 2196 cases, HoNOS was used in 800 cases, neither was available in 62 cases.  - mild=MMSE>22 or HONOS=0-2  - mod=MMSE 18-22 or HONOS=3  - severe=MMSE<18 or HONOS=4  - unknown=no MMSE or HONOS in window |
| Activities of daily living problems | CRIS – Mentalhealthcare full text | closest to index | Including moderate or severe problem (score 3 or 4) |
| Health |  |  |  |
| History of falls | LDN – Primary care data | up until index | From Caliber Read code list |
| GP contact | LDN – Primary care data | two years before index | A count for face-to-face and phone/virtual consultations, split into above and below median |

AD = Alzheimer’s disease; AEC = Anticholinergic effect on cognition; CRIS = Clinical Research Information Search (at South London and Maudsley); HoNOS = Health of the Nation Outcome Scales (65+); LDN = Lambeth DataNet; MMSE = Mini-Mental State Examination

S3 Supplementary Table 3**. Baseline demographic, clinical and comorbidity characteristics of the dementia cohort (n = 3,543)**

| **Characteristic** | **State** | **n** | **% cohort** |
| --- | --- | --- | --- |
| **Age at diagnosis (years)** | 65–69 | 222 | 6% |
|  | 70–74 | 394 | 11% |
|  | 75–79 | 679 | 19% |
|  | 80–84 | 866 | 24% |
|  | 85–89 | 851 | 24% |
|  | 90–94 | 394 | 11% |
|  | ≥95 | 137 | 4% |
| **Sex** | Female | 2,076 | 59% |
|  | Male | 1,467 | 41% |
| **Index of Multiple Deprivation (IMD)** | Below average | 1,741 | 49% |
|  | Average | 1,098 | 31% |
|  | Above average | 704 | 20% |
| **Marital status** | Married/cohabiting | 898 | 25% |
|  | Other | 2,054 | 58% |
|  | Missing | 591 | 17% |
| **Ethnicity** | White | 2,044 | 58% |
|  | Black | 933 | 26% |
|  | Other | 356 | 10% |
|  | Unknown | 210 | 6% |
| **Dementia severity** | Mild | 956 | 27% |
|  | Moderate | 736 | 21% |
|  | Severe | 1,315 | 37% |
|  | Unknown | 536 | 15% |
| **Dementia subtype** | Alzheimer's disease | 2,242 | 63% |
|  | Vascular/mixed | 646 | 18% |
|  | Other | 167 | 5% |
|  | Unknown | 488 | 14% |

S4 Supplementary Table 4: Detail of internal validation of MedCAT comorbidities at instance level

| **Condition** | n. instances examined | Precis-ion | False positives (all causes of >2 per topic) |
| --- | --- | --- | --- |
| Liver | 14 | 100% | Nil |
| Atrial Fibrillation | 213 | 99% | Nil |
| Myocardial infarction | 333 | 99% | someone else (4) |
| Renal | 231 | 98% | Nil |
| Hypertension | 199 | 95% | form (3), referring to a BP reading (3) |
| COPD | 335 | 94% | form (8), refers to other (5), suspected (3) |
| Coronary arteriosclerosis | 156 | 92% | risk score (8), at risk (3) |
| Cerebrovascular accident | 343 | 91% | suspected (20), form (4) |
| TIA | 170 | 90% | suspected (12), form (5) |
| Ischaemic HD | 215 | 89% | resolved angina (13), refers to other (4), suspected (4) |
| Heart failure | 150 | 89% | suspected (7), form (5) |
| Diabetes | 349 | 84% | form (26), pre-diabetes and borderline diabetes (15), at risk (4), refers to other (3) |

S5 Supplementary Table 5 The 10 most common comorbid conditions in the cohort (n=3543) as ascertained by the individual data sources, and in combination: Clinical Record Interactive Search (CRIS) full text, via MedCAT comorbidity app, Lambeth DataNet (LDN) via codelist, Hospital Episode Statistics (HES) via codelist

| *Any of MedCAT, primary or hospital* |  | MedCAT NLP in mental health text |  | Primary care data |  | Hospital inpatient data |  |
| --- | --- | --- | --- | --- | --- | --- | --- |
| Hypertension | 82% | Hypertension | 64% | Hypertension | 58% | Hypertension | 70% |
| Diabetes | 51% | Diabetes | 42% | Diabetes | 37% | Stroke/TIA | 37% |
| Stroke/TIA | 50% | Stroke/TIA | 27% | Renal | 32% | Diabetes | 34% |
| Renal | 39% | Renal | 22% | Stroke/TIA | 24% | AF | 27% |
| Respiratory | 33% | Respiratory | 20% | Respiratory | 21% | Respiratory | 25% |
| AF | 30% | Coronary HD | 15% | AF | 19% | Renal | 23% |
| Heart Failure | 25% | AF | 13% | Heart failure | 12% | Heart Failure | 19% |
| Coronary HD | 22% | Heart failure | 11% | Coronary HD | 10% | MI | 13% |
| Neuro | 17% | Neuro | 10% | MI | 8% | Coronary HD | 12% |
| MI | 15% | MI | 6% | Neuro | 7% | Neuro | 12% |

AF = Atrial Fibrillation, Coronary HD = Coronary Heart Disease, MI = Myocardial Infarction, Neuro = Neurological, TIA = Transient Ischaemic Attack

S6 Supplementary Table 6 **The RECORD statement – checklist of items, extended from the STROBE statement, that should be reported in observational studies using routinely collected health data.**

|  | **Item No.** | **STROBE items** | **Location in manuscript where items are reported** | **RECORD items** | **Location in manuscript where items are reported** |
| --- | --- | --- | --- | --- | --- |
| **Title and abstract** | | | | | |
|  | 1 | (a) Indicate the study’s design with a commonly used term in the title or the abstract (b) Provide in the abstract an informative and balanced summary of what was done and what was found | Title/Abstract – validation study; informative summary of methods and findings. | RECORD 1.1: The type of data used should be specified in the title or abstract. When possible, the name of the databases used should be included.  RECORD 1.2: If applicable, the geographic region and timeframe within which the study took place should be reported in the title or abstract.  RECORD 1.3: If linkage between databases was conducted for the study, this should be clearly stated in the title or abstract. | RECORD 1.1 – EHR stated in title/abstract; CRIS, LDN and HES named in the abstract/methods.  RECORD 1.2 – South London stated in the abstract; study timeframe is not stated in the title/abstract.  RECORD 1.3 – Linkage stated in the title and abstract. |
| **Introduction** | | | | | |
| Background rationale | 2 | Explain the scientific background and rationale for the investigation being reported | Y – Introduction |  |  |
| Objectives | 3 | State specific objectives, including any prespecified hypotheses | Y – Introduction, objectives (1)–(3) |  |  |
| **Methods** | | | | | |
| Study Design | 4 | Present key elements of study design early in the paper | Y – Methods, Data Sources/Cohort/Analyses 1–3 |  |  |
| Setting | 5 | Describe the setting, locations, and relevant dates, including periods of recruitment, exposure, follow-up, and data collection | Y – Methods, Data Sources/Cohort; extraction 02/11/2022, HES to 31/03/2022; four-year mortality follow-up |  |  |
| Participants | 6 | *(a) Cohort study* - Give the eligibility criteria, and the sources and methods of selection of participants. Describe methods of follow-up  *Case-control study* - Give the eligibility criteria, and the sources and methods of case ascertainment and control selection. Give the rationale for the choice of cases and controls  *Cross-sectional study* - Give the eligibility criteria, and the sources and methods of selection of participants  *(b) Cohort study* - For matched studies, give matching criteria and number of exposed and unexposed  *Case-control study* - For matched studies, give matching criteria and the number of controls per case | Y – Methods, Cohort (eligibility, data sources, exclusions and follow-up window) | RECORD 6.1: The methods of study population selection (such as codes or algorithms used to identify subjects) should be listed in detail. If this is not possible, an explanation should be provided.  RECORD 6.2: Any validation studies of the codes or algorithms used to select the population should be referenced. If validation was conducted for this study and not published elsewhere, detailed methods and results should be provided.  RECORD 6.3: If the study involved linkage of databases, consider use of a flow diagram or other graphical display to demonstrate the data linkage process, including the number of individuals with linked data at each stage. | RECORD 6.1 – Y, Methods Cohort + Supplementary Code Lists  RECORD 6.2 – Y, SAIL/CALIBER algorithms referenced  RECORD 6.3 – Linkage described and referenced; no flow diagram |
| Variables | 7 | Clearly define all outcomes, exposures, predictors, potential confounders, and effect modifiers. Give diagnostic criteria, if applicable. | Y – Methods, Defining conditions/Other variables/Analysis 3 | RECORD 7.1: A complete list of codes and algorithms used to classify exposures, outcomes, confounders, and effect modifiers should be provided. If these cannot be reported, an explanation should be provided. | Y – Supplementary Code Lists; Supplementary Tables S1–S2 |
| Data sources/ measurement | 8 | For each variable of interest, give sources of data and details of methods of assessment (measurement).  Describe comparability of assessment methods if there is more than one group | Y – Methods, Data Sources and Defining conditions; Supplementary Tables S1–S2 |  |  |
| Bias | 9 | Describe any efforts to address potential sources of bias | Y – Analyses 1–3 (internal validation, external cross-validation and predictive validity); limitations discuss misclassification/under-documentation |  |  |
| Study size | 10 | Explain how the study size was arrived at | Y – available cohort size after eligibility determine study size without prior power calculation; final n=3,543 in Results |  |  |
| Quantitative variables | 11 | Explain how quantitative variables were handled in the analyses. If applicable, describe which groupings were chosen, and why | Y – see analyses, no groupings other than described in S1 and S2 |  |  |
| Statistical methods | 12 | (a) Describe all statistical methods, including those used to control for confounding  (b) Describe any methods used to examine subgroups and interactions  (c) Explain how missing data were addressed  (d) *Cohort study* - If applicable, explain how loss to follow-up was addressed  *Case-control study* - If applicable, explain how matching of cases and controls was addressed  *Cross-sectional study* - If applicable, describe analytical methods taking account of sampling strategy  (e) Describe any sensitivity analyses | Y – (a) Methods, Analyses 1–3 describe precision/recall, Cox regression, confounder adjustment and Schoenfeld residuals.  (b) none  (c) known missing data described in other variables. |  |  |
| Data access and cleaning methods |  | .. |  | RECORD 12.1: Authors should describe the extent to which the investigators had access to the database population used to create the study population.  RECORD 12.2: Authors should provide information on the data leaning methods used in the study. | RECORD 12.1 – Methods, Ethics and data availability.  RECORD 12.2 – Data-cleaning via rstudio |
| Linkage |  | .. |  | RECORD 12.3: State whether the study included person-level, institutional-level, or other data linkage across two or more databases. The methods of linkage and methods of linkage quality evaluation should be provided. | Y – stated person-level. Papers provided for greater detail |
| **Results** | | | | | |
| Participants | 13 | (a) Report the numbers of individuals at each stage of the study (*e.g.*, numbers potentially eligible, examined for eligibility, confirmed eligible, included in the study, completing follow-up, and analysed)  (b) Give reasons for non-participation at each stage.  (c) Consider use of a flow diagram | Partial – Results, Cohort reports last step of selection n=3,543. Representativeness less relevant for study. | RECORD 13.1: Describe in detail the selection of the persons included in the study (*i.e.,* study population selection) including filtering based on data quality, data availability and linkage. The selection of included persons can be described in the text and/or by means of the study flow diagram. | Partial – Methods, Cohort describes filtering by GP registration, CRIS window and opt-out due to data availability. Non-included are not known due to data extraction methods |
| Descriptive data | 14 | (a) Give characteristics of study participants (*e.g.*, demographic, clinical, social) and information on exposures and potential confounders  (b) Indicate the number of participants with missing data for each variable of interest  (c) *Cohort study* - summarise follow-up time (*e.g.*, average and total amount) | Y – Results, Cohort and Supplementary Table S3 report demographic/clinical characteristics, including missing data ‘unknown’. Four-year follow-up is used for survival analysis, and since death is assumed to be reliably ascertained, follow-up assumed to be complete. |  |  |
| Outcome data | 15 | *Cohort study* - Report numbers of outcome events or summary measures over time  *Case-control study* - Report numbers in each exposure category, or summary measures of exposure  *Cross-sectional study* - Report numbers of outcome events or summary measures | Y – Results, Modelling comorbidities and mortality: 43% mortality during four-year follow-up; Table 5 reports mortality model outcomes. |  |  |
| Main results | 16 | (a) Give unadjusted estimates and, if applicable, confounder-adjusted estimates and their precision (e.g., 95% confidence interval). Make clear which confounders were adjusted for and why they were included  (b) Report category boundaries when continuous variables were categorized  (c) If relevant, consider translating estimates of relative risk into absolute risk for a meaningful time period | Partial – Table 5 reports adjusted HRs and p-values; adjustment variables are specified in Methods/table footnote. Unadjusted estimates and 95% CIs are not reported. |  |  |
| Other analyses | 17 | Report other analyses done—e.g., analyses of subgroups and interactions, and sensitivity analyses | NA – no subgroup, interaction or sensitivity analyses are reported. |  |  |
| **Discussion** | | | | | |
| Key results | 18 | Summarise key results with reference to study objectives | Y – Discussion opening paragraph summarises findings against the three stated aims; Conclusion reiterates the key results. |  |  |
| Limitations | 19 | Discuss limitations of the study, taking into account sources of potential bias or imprecision. Discuss both direction and magnitude of any potential bias | Y – Discussion and Strengths and Limitations; under-ascertainment, misclassification and uncertainty from low-prevalence conditions are discussed. | RECORD 19.1: Discuss the implications of using data that were not created or collected to answer the specific research question(s). Include discussion of misclassification bias, unmeasured confounding, missing data, and changing eligibility over time, as they pertain to the study being reported. | Y – Discussion addresses routinely collected data, under-documentation, misclassification and non-random missingness. Unmeasured confounding and changing eligibility not relevant |
| Interpretation | 20 | Give a cautious overall interpretation of results considering objectives, limitations, multiplicity of analyses, results from similar studies, and other relevant evidence | Y – Discussion and Conclusion; cautious interpretation considers limitations and results from previous studies. |  |  |
| Generalisability | 21 | Discuss the generalisability (external validity) of the study results | Y – Strengths and Limitations: single geographical area may limit generalisability, despite an ethnically diverse population. |  |  |
| **Other Information** | | | | | |
| Funding | 22 | Give the source of funding and the role of the funders for the present study and, if applicable, for the original study on which the present article is based | N -Not reported in the current manuscript |  |  |
| Accessibility of protocol, raw data, and programming code |  | .. |  | RECORD 22.1: Authors should provide information on how to access any supplemental information such as the study protocol, raw data, or programming code. | Partial – Supplementary Code Lists are referenced. Raw data statement given. |

*Reference: Benchimol EI, Smeeth L, Guttmann A, Harron K, Moher D, Petersen I, Sørensen HT, von Elm E, Langan SM, the RECORD Working Committee. The REporting of studies Conducted using Observational Routinely-collected health Data (RECORD) Statement. *PLoS Medicine* 2015; in press.

*Checklist is protected under Creative Commons Attribution ([CC BY](http://creativecommons.org/licenses/by/4.0/)) license.
